# The impact of threshold decision mechanisms of collective behaviour on disease spread

**DOI:** 10.1101/2022.11.22.22282606

**Authors:** Bryce Morsky, Felicia Magpantay, Troy Day, Erol Akçay

## Abstract

Humans are a hyper social species, which greatly impacts the spread of infectious diseases. How do social dynamics impact epidemiology? How does public health policy best take into account these impacts? Here we develop a model of disease transmission that incorporates human behaviour and social dynamics. We use a “tipping-point” dynamic, previously used in the sociological literature, where individuals adopt a behaviour given a sufficient frequency of the behaviour in the population. The thresholds at which individuals adopt behaviours is modulated by the perceived risks of infection, i.e. the disease prevalence and transmission rate, and the behaviour of others. Social conformity creates a type of “stickiness” whereby individuals are resistant to changing their behaviour due to the population’s inertia. In this model, the epidemic attack rate is sensitive to the timing of the behavioural response. Near the optimal response, small errors can result in large increases in the total number infected during the epidemic. And, more surprisingly, we observe a non-monotinicity in the attack rate as a function of various biological and social parameters such as the transmission rate, efficacy of social distancing, the costs to social distancing, the weight of social consequences of shirking the norm, and the degree of heterogeneity in the population.

## 1 Introduction

The spread of pathogens in human populations crucially depends on social, political, psychological, and economic factors (Soofi et al., 2020). Informal social rules such as norms and cultural practices can impact the efficacy of treatment and public policy through their effects on human behaviour. Such behavioural factors may either promote or inhibit the spread of disease. The COVID-19 pandemic has demonstrated with unusual force how critical the interactions between social and epidemiological dynamics are to controlling diseases, and how much we still have to learn about them.

There is a rich literature of disease models incorporating social behaviour (Jansen et al., 2003; Funk et al., 2010). Some models consider behavioral change in response to the disease as a parallel epidemic where behavior is spread through contact (Funk et al., 2009; Smaldino and Jones, 2021), whereas others model behavioral change with a game theoretic analysis. Game theory models tend to focus on if and when individually optimal behavior leads to optimality of aggregate outcomes for social welfare. A common intuition from game theory (and deeply ingrained in many cultures) is that selfish motives by the players can lead to suboptimal outcomes for a society. Indeed, this is the focus of much literature on cooperation faced with a social dilemma. For example, in the context of infectious diseases, a social dilemma exists with respect to vaccination, where a population can be prevented from reaching herd immunity (Bauch et al., 2003). Other game-theoretic examples consider the impact of self-interested individual decision-making on disease dynamics, where individuals weigh risks and rewards to determine their optimal strategies for reducing transmission risk (Morsky and Bauch, 2012; Fenichel, 2013).

In addition to economic and risk incentives, human behaviour with respect to disease spread is strongly shaped by socially transmitted behaviours and social norms, the unwritten rules of social interactions (Bicchieri, 2005). Social norms help determine the expectations individuals have on both what others will do and what they should do, and thus guide personal decision making. In the context of disease spread, socially transmitted practices among doctors and expectations amongst patients contributed to the emergence of antibiotic resistance due to antibiotic overuse (McGowan Jr, 1983; Austin et al., 1999; Karakonstantis and Kalemaki, 2019). Likewise, West African burial traditions contributed to the spread of Ebola (Manguvo and Mafuvadze, 2015; Alexander et al., 2015), and anti-vaccination movements contributed to the spread of measles (Patricia et al., 2019). On the other hand, social norms can be helpful in preventing the spread of disease, such as in East Asia where social acceptance of mask wearing is high (Liu, 2021). Mask wearing can be driven by not only belief in its effectiveness but also in the prevalence of mask wearing (Bokemper et al., 2021). Further, adherence to norms of mask wearing can vary within a population due to group identity, which can frustrate behavioural interventions (Cakanlar et al., 2022; Dimant et al., 2022; Gelfand et al., 2022). Though norms can lead to coordination of human behaviour (Gintis, 2010; Morsky and Akçay, 2019) and the emergence of cooperative communities (Ostrom, 2000; Morsky and Akçay, 2021) thereby overcoming social dilemmas, they also can lead to harmful outcomes and be difficult to dislodge (Efferson et al., 2020). It is important to incorporate these social phenomena into epidemiological theory and the design of public health control efforts.

Here we aim to understand how social dynamics drive the use of non-pharmaceutical interventions (NPI), specifically social distancing, in blunting the spread of disease. Previous studies have looked at how a rational actor would behave in epidemics (Gersovitz and Hammer, 2003; Toxvaerd, 2020; Bhattacharya et al., 2021). However, social dynamics driven by imitation and social norms are equally, if not more important, than decision making of rational actors, and there is a growing literature incorporating them (Weitz et al., 2020; Qiu et al., 2022). We develop an epidemiological model that incorporates social dynamics of NPI usage where individuals weigh the risks of infection, the cost of using the NPI, and the social cost of diverging from a social norm for the NPI usage. We do this by combining material and relational utility and applying a threshold model of collective decision making. The model induces a “tipping-point” dynamic, introduced in social behaviour models (Schelling, 1969, 1971; Granovetter, 1978; Granovetter and Soong, 1983, 1986, 1988), and which has been found in other models of disease spread (Qiu et al., 2022). We also observe non-monotonicity in the attack rate as a function of the transmission rate but show that it can be more complex than a single shift down. Additionally, we oberserve non-monotonicity when changing the efficacy of the NPI and the costs individuals face in their decision-making. From these observations, we find that an intermediate level for a variety of key parameters can be optimal in reducing the attack. However, the optimal levels are frequently highly sensitive to changes in parameters.

## 2 Methods

We consider the classic SEIR model with the addition of a behavioural dynamic. Our equations are:

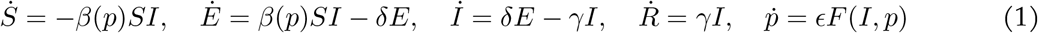

with *S, E, I*, and *R* the frequency of susceptible, exposed, infectious, and recovered individuals, and *p* ∈ [0, 1] the degree to which the susceptible population adopt the NPI. The quantities 1*/δ* and 1*/γ* are the mean latent and recovery periods respectively. We assume that *p* affects the transmission rate *β*(*p*). Specifically, *β*(*p*) is a non-increasing function where *β*(0) = *β*_0_, the transmission rate without NPI usage. There is evidence from mask mandates of risk compensation (Yan et al., 2021). However, we assume that any risk compensation due to a higher efficacy of the NPI is not sufficient to increase the transmission rate. We will primarily consider the case where *β*(*p*) = *β*_0_(1 − *ηp*) with *η* ∈ [0, 1] being the efficacy of the NPI in reducing transmission. We also considered *β*(*p*) = *β*_0_*/*(1 + *ηp*) with *η* ≥ 0, which showed no great qualitative difference in the outcome (results not shown here). For numerical simulations, the standard parameter values we use are displayed in Table 1.

**Table 1:** Summary definitions of parameters with default values.

| Parameter | Definition | Default value |
| --- | --- | --- |
| $\beta_0$ | transmission rate without the NPI | 0.4/day |
| $1/\gamma$ | average recovery period | 10 days |
| $1/\delta$ | average latency period | 5 days |
| $\epsilon$ | behavioural change rate | 1/day |
| $\eta$ | efficacy of the NPI | 0.8 |
| $\kappa$ | payoff differential sensitivity | 1000 |
| $c$ | material cost | 0.02 |
| $\theta$ | relational cost | 0.01 |

The rate at which the transmission affecting behaviour changes, *dp/dt* is governed by *F* (*I, p*), which is a function of the current number of infectious individuals, the current behaviour of the population, *p* (which we assume individuals know). The parameter *ϵ* governs the magnitude of the rate of change of *p*. These assumptions are reasonable in the case of conspicuous NPI’s or rapid gossip. We consider a Granovetter-Schelling updating process for the adoption of the NPI (Schelling, 1971; Granovetter and Soong, 1983), which is a well known dynamic in the social science literature for threshold action. The equation for this dynamic is:

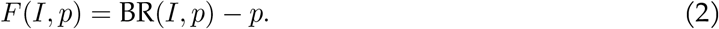

BR is a smoothed “best response” function to the current perceived state of the epidemic:

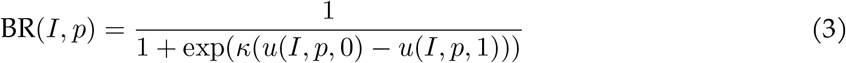

where *u*(*I, p, q*) is the individual utility to adopt the NPI with probability *q* when the density of infections is *I* and the overall fraction of susceptible individuals that adopt the NPI is *p*. Thus *u*(*I, p*, 1) is the utility for an individual fully compliant with the NPI, and *u*(*I, p*, 0) is the utility to an individual who has not adopted it. Individuals compare these two payoffs to determine their behaviour. The constant *κ >* 0 is the sensitivity to the payoff differential. Thus, for example, if *κ* is large, then the best response for an individual is to adopt the NPI if *u*(*I, p*, 0) *< u*(*I, p*, 1) and not to adopt it if *u*(*I, p*, 0) *> u*(*I, p*, 1). By Equation 2, *p* increases in the former case, and decreases in the latter.

The utility function is a sum of material (economic) utility *u*^*m*^(*I, q*) and relational (social) utility *u*^*r*^(*p, q*). Material utility is a function of the risk of infection (related to the number infectious individuals and the degree of social distancing) and the economic cost of social distancing. We set *u*^*m*^(*I, q*) = −*β*(*q*)*I* − *cq* so that it is a decreasing function with respect to *I*. It is increasing or decreasing with respect to *q* dependent on the epidemic state of the population. When infections are low, the NPI is primarily costly due to the cost *c* to adopt it. When infections are high, the reduced chance of infection from the NPI outweighs this cost, and thus material utility increases with respect to *q*. On the other hand, relational utility is solely governed by what individuals do and what others do. Guilt and pressure to socially conform can be relational utilities and can overwhelm economic incentives. Under a promoting norm (Su and Morsky, 2022), relational utility favours a specific behaviour — such as NPI usage — and is relative to the average behaviour in the population. In such a case, one earns positive relational utility (a “warm glow”) if *q > p*, and one earns negative utility (guilt, social pressure) (Battigalli and Dufwenberg, 2007) if *q < p*. Such a norm can lead to bistability, however, if the initial adoption of the norm is low (Su and Morsky, 2022). The norm that we focus on here is a norm of conformity in which deviations from the population average impose negative relational utility (Akçay and Van Cleve, 2019; Su and Morsky, 2022). Under this norm *u*^*r*^(*p, q*) = −*θ*(*p* − *q*)^2^. The relational utility is related to the average behaviour in the population and is mediated by the parameter *θ*, which is a weighting of the cost to deviating from the norm. Utility is thus given by,

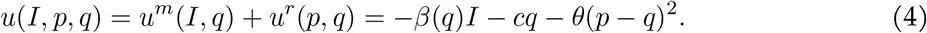

As the epidemic spreads, these utilities have an impact on the threshold at which the NPI is adopted.

## 3 Results

According to our best-response function and assuming *κ* ≫ 1, individuals should start favoring the NPI behavior if *u*(*I, p*, 1) *> u*(*I, p*, 0) which occurs when,

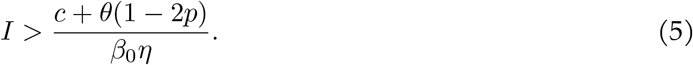

This threshold is a function of both the material and social utilities. On the material side, the threshold decreases as the efficacy of the norm *η* increases, and increases as its cost *c* increases. On the social side, the relational utility cost *θ* has divergent effects depending on population behavior: if the majority of the population adopts the NPI (i.e., *p >* 1*/*2), then the threshold decreases as *θ* increases. However, if only a minority adopt the NPI (i.e., *p <* 1*/*2), then increasing the relational cost increases the threshold. Social conformity can thus retard initial adoption of the NPI relative to when it would be materially rational to adopt it. On the other hand, it also causes individuals to use the NPI longer than they should with respect to material well-being. This norm “stickiness” is illustrated in Figure 1a. As the number of infectious individuals increases, the best response function shifts, which can shift equilibrium behaviour from a state where only a few individuals adopt the NPI to a bistable system where most people will either adopt the NPI or not, depending on what they believe others do, and then to a state where almost all engage in the behaviour. Such phenomena have been previously explored in the formation and dynamics of social groups (Morsky and Akçay, 2021). Figures 1b and 1c depict such shifts.

**Figure 1:**
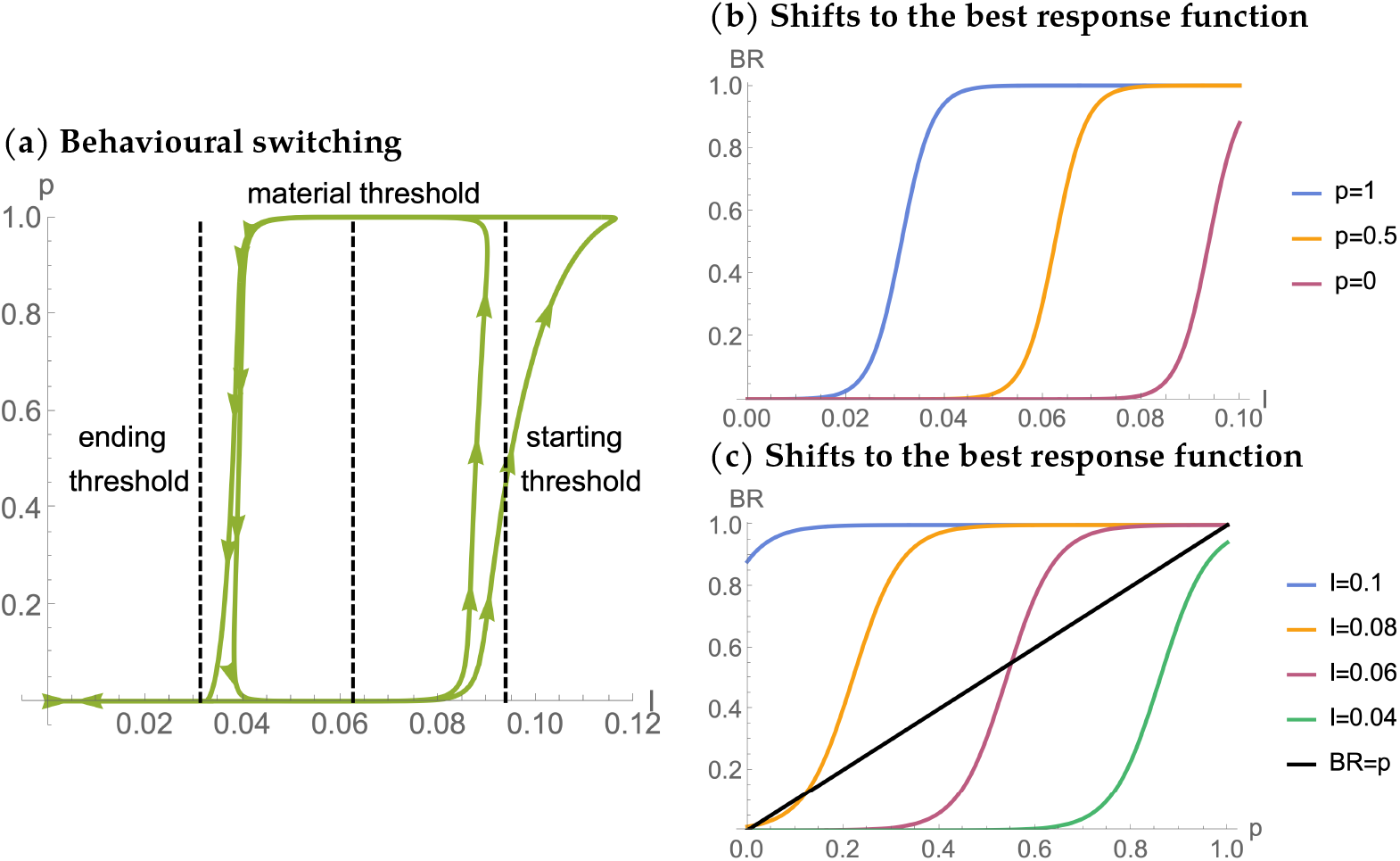
Coupled dynamics of behavioral change and epidemic caused by behavioral switches induced by norms. (a) Sample trajectory of an epidemic depicted in the space of infectious individuals *I* and fraction of NPI adopters *p*. The arrows depict the direction of the trajectory, which is counter-clockwise. The dashed vertical lines give different thresholds for switching behavior (i.e., the infection levels when the best response function favors adoption or abandonment of the NPI, equation (5)). When there is no relational utility (no social norms or *θ* = 0), the thresholds for adoption and abandonment are the same (depicted by the “material threshold” in the middle). With relational utility (*θ >* 0), the adoption of the behavior happens at higher infection levels (“starting threshold”) but abandonment at lower infection levels compared to the material threshold. Panels b and c show how the best response curve shifts when varying *p* and *I*. Panel b shows increasing *p* shifts the best response curve in *I* to the left, i.e., favors adoption of the behavior for the individual for a given infection level *I*. Likewise, Panel c shows that increasing *I* shifts the best response curve in *p* to the left; favoring adoption for a given fraction of NPI adopters. The intersection of the diagonal line in panel c and the best response curve is when 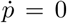. When *I* is low, the system has two stable fixed points in *p* (high and low) and an intermediate, unstable one. As *I* increases, the intermediate unstable equilibrium disappears and the system shifts to the high *p* equilibrium. The shifting of the curve The parameter values are taken from Table 1.

The switching dynamic can produce behavioural waves of NPI usage, which in turn can generate epidemic waves: Figures 2b-2e are examples. As the frequency of infectious individuals dips above and below the threshold of equation 5, NPI’s are rapidly heavily employed and then rapidly abandoned. The number of these waves are attenuated by the parameters. As shown in Figures 2b-2e, a higher efficacy of the NPI can produce further waves. The lower the threshold of infectious individuals, the more waves can be induced. Regardless of the number of waves in which behavior change is induced, there is always an “exit wave” where infections never again rise to the level required to trigger the behavior change. The size of this exit wave is a function of the number of susceptibles left after the last wave with behavioural change abates.

**Figure 2:**
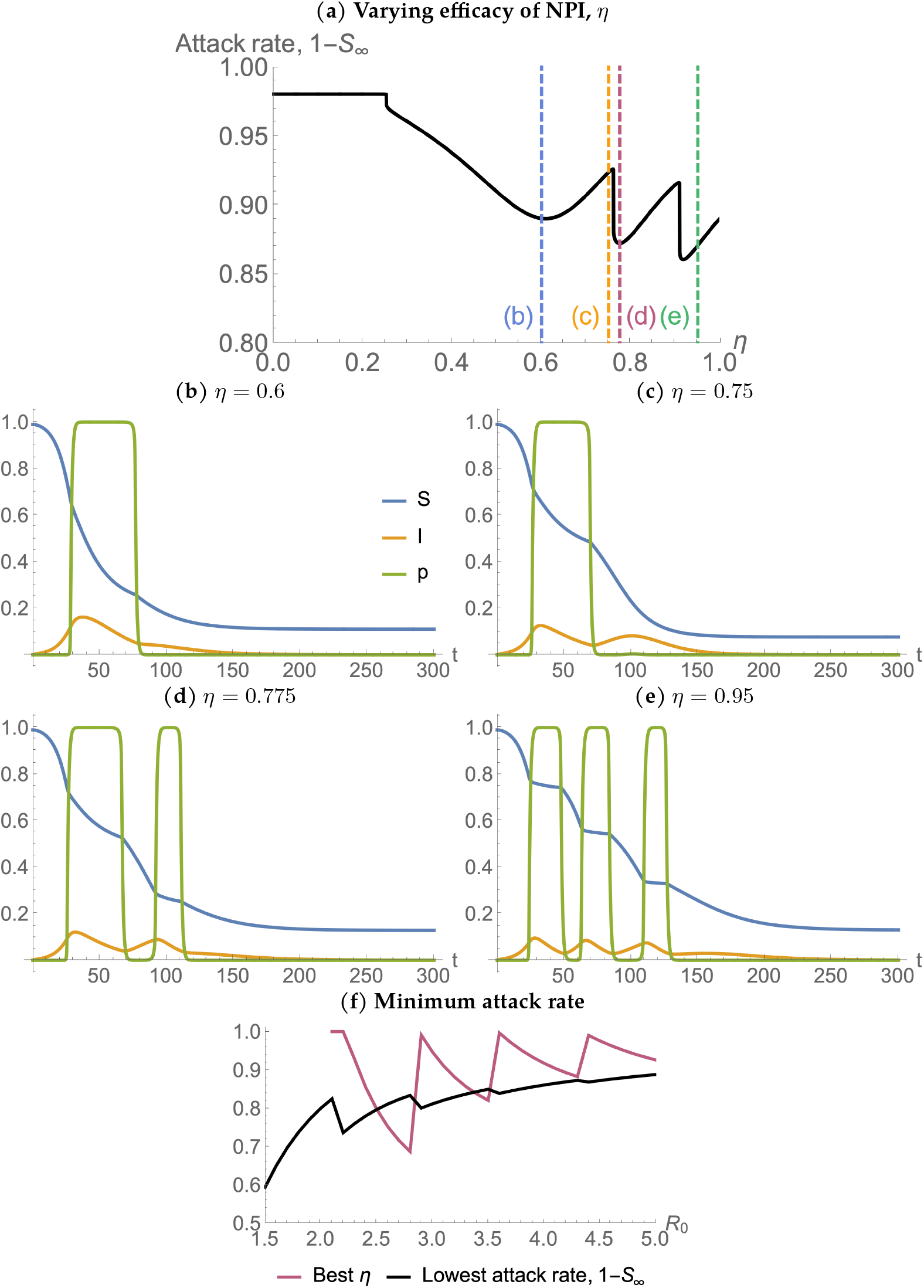
The attack rate changes non-monotonically with changing *η*, the efficacy of the NPI behavior. Panel a shows that there is a saw-tooth pattern in *η* and increasing number of behaviour waves as *η* increases. Panels b-e depict the time trajectories of the epidemic and behavioral change for different values of *η*, as marked by the vertical dashed lines on Panel a. Panel (f) depicts the minimum attack rate (black) and the NPI efficacy *η* needed to obtain it as a function of *R*_0_ (varied by changing *β*_0_). Unless otherwise stated, the parameter values are taken from Table 1.

The number of waves and the impact of parameters can have counter-intuitive effects on the attack rate of the epidemic. We find a non-monotonicity in the attack rate when varying the parameters within the threshold equation 5. Figure 2a depicts the attack rate when we vary the material and relational costs. We can explain this result by noting that there is a trade-off when NPI efficacy is increased. The higher the NPI efficacy, the earlier the population reacts to the epidemic, since the threshold to adopt the behaviour is lower. Although the threshold to end the behaviour is also lower, the time until this threshold is reached can be longer or shorter. Thus, the duration of NPI usage does not depend monotonically upon the NPI efficacy. Likewise, increasing NPI efficacy also decreases the transmission rate during the period of social distancing. Both of these effects lead to a larger pool of susceptibles immediately prior to the exit wave, and thereby a larger exit wave. We can develop some mathematical intuition of this by considering an SIR dynamic (where *p* = 0 and does not change) and examining the final size equation (Murray, 2002)

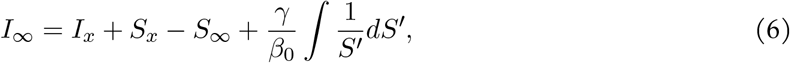

where *S*_∞_ and *I*_∞_ = 0 are the final sizes of susceptibles and infectious, and *S*_*x*_ and *I*_*x*_ are the initial sizes at the beginning of the exit wave. Here we’re assuming that the NPI was successful in suppressing the infection to low levels, and hence *I*_*x*_ ≈ 0. Rearranging we thus have

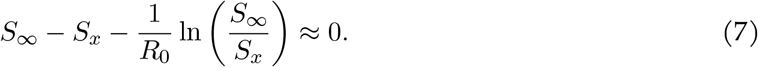

Differentiating with respect to *S*_*x*_ provides:

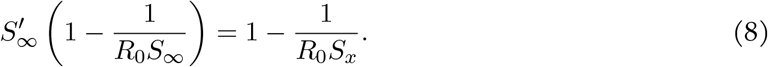

Assuming there is an exit wave, we have *R*_0_*S*_*x*_ *>* 1 and *R*_0_*S*_∞_ *<* 1. Therefore the attack rate 1 − *S*_∞_ is increasing with respect to *S*_*x*_. Lowering the threshold increases the duration of NPI use, and increases the number of susceptibles left at the end of the wave. Therefore increasing efficacy may increase the size of the exit wave and thus the attack rate. However, increasing effectiveness further can lower the threshold infection numbers such that what previously was an exit wave triggers another wave of behavioral change. This would cause a drop in *S*_*x*_ such that the final epidemic size drops again. This effect is what causes the saw-tooth pattern in Figure 2a in the final epidemic size. Thus, there is a trade-off in increasing the NPI efficacy, which results in an intermediate level of NPI efficacy being (locally) optimal. Further, this optimal efficacy varies with respect to *R*_0_. Figure 2f depicts the best NPI efficacy for different *R*_0_. Below a threshold *R*_0_ ≈ 2, behavioural waves are never initiated. Above this threshold, however, a saw-tooth pattern emerges: the sharp changes of which make policy recommendations that impact NPI efficacy difficult. Given some uncertainty on *R*_0_, it could be difficult to determine the optimal NPI efficacy needed. See Appendix A.1 for further mathematical details of the impact of *η* on the attack rate.

Varying the intrinsic transmission rate *β*_0_ — the rate when there is no NPI usage — also results in a non-monotonic impact on the attack rate as shown in Figure 3a. For sufficiently low transmission rates, no behavioural wave is induced (Figure 3b). Once transmission is sufficient to induce such a wave, we see a drop in the attack rate (Figure 3c). A further increase results in another wave and thereby another drop in the attack rate (Figure 3d). However, further increasing the transmission rate can reduce the number of behavioural waves and thereby increase the attack rate. The higher transmission rate results in an earlier onset of the behavioural wave. However, it also increases its duration, since the number of infectious individuals remains sufficiently high — due to the higher transmission rate — to promote NPI usage. This effect causes the secondary wave of infections to not induce a secondary behavioural wave. Figure 3e typifies this situation. See Appendix A.2 for further mathematical analysis.

**Figure 3:**
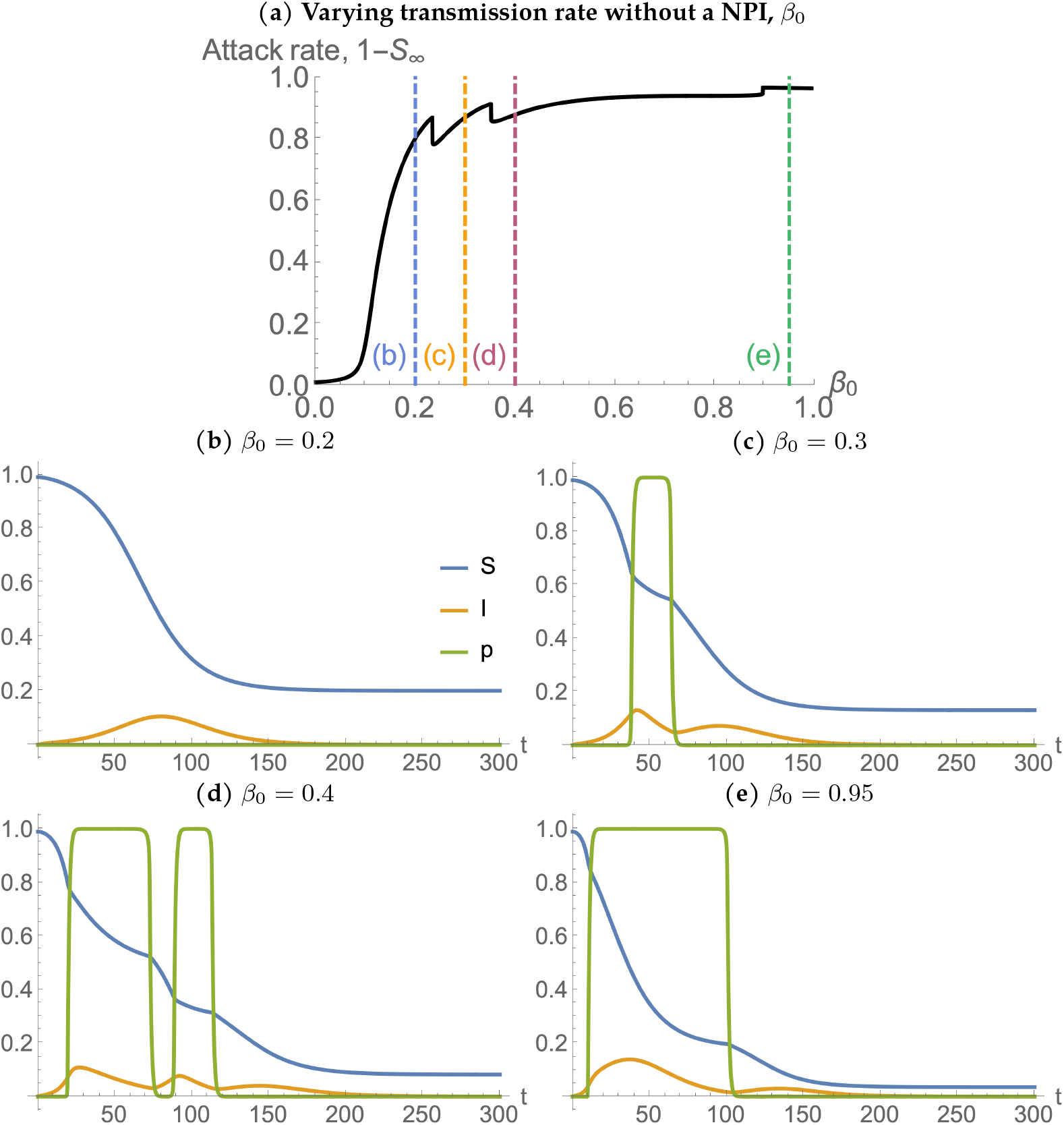
The attack rate changes non-monotonically with changing baseline transmission rate, *β*_0_. As in Figure 2a, Panel a depicts a saw-tooth pattern of the attack rate as *β*_0_ varies, and panels b-e depict time trajectories for different values of *β*_0_, as marked by vertical dashed lines in panel a. For increasing *β*_0_, the number of behavioural waves goes from zero, to one, to two, and back to one. Unless otherwise stated, the parameter values are taken from Table 1.

Increasing the material cost *c* can reduce the number of waves, which explains the jumps we observe in the attack rate in Figure 4a. Figures 4b–4e depict time series for different values of *c*. Jumps in the attack rate can occur due to changes in the number of waves. However, increasing the cost will only increase the attack rate assuming that there is only one behavioural response wave (see Appendix A.3 for mathematical details). Another observation of the impact of varying material costs is that having a low *c* extends the pandemic, since people adopt the NPI as soon as there are flare-ups. For example, it takes longer for the pandemic to be resolved in Figure 4c than it does in Figures 4d and 4e.

**Figure 4:**
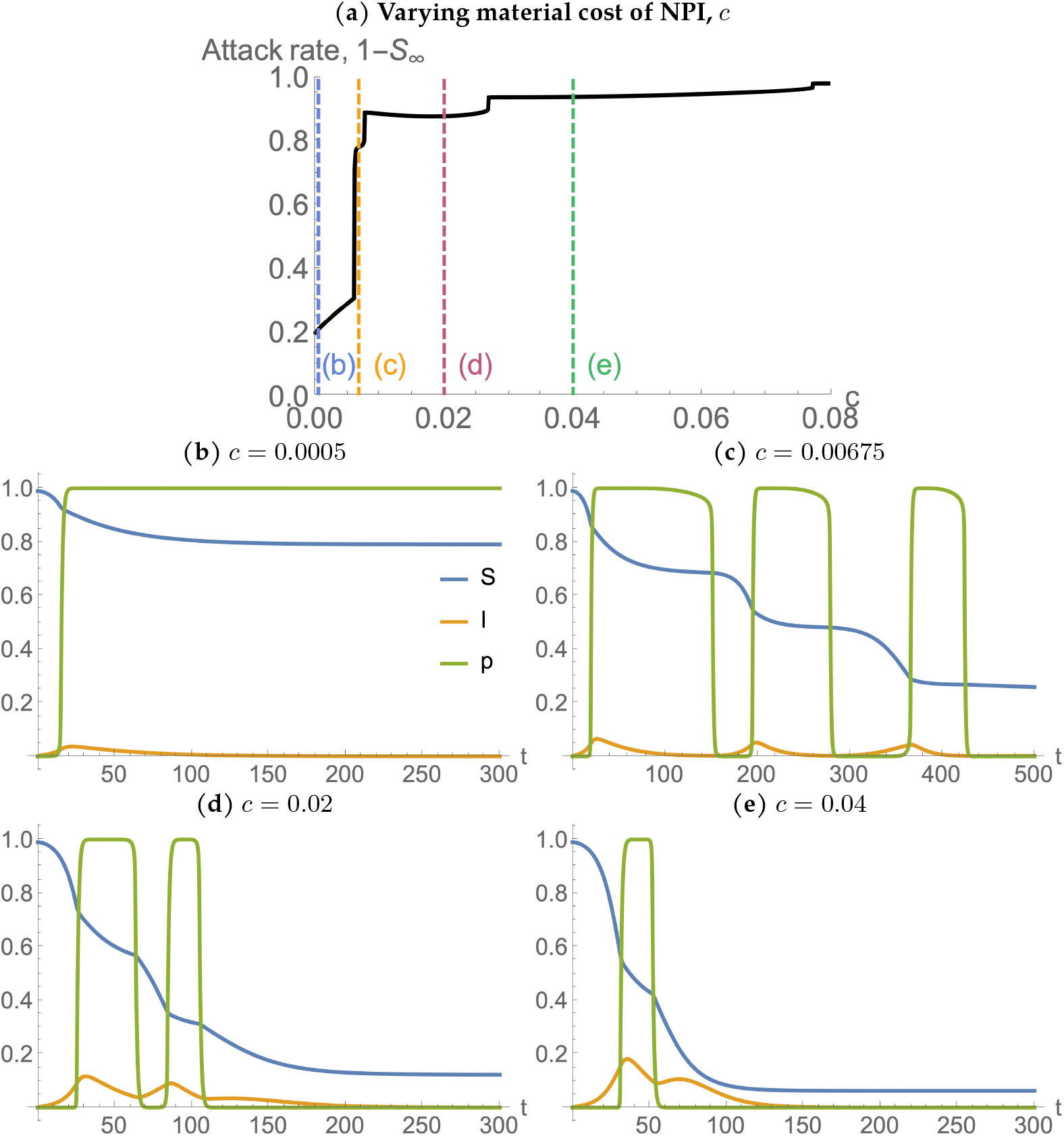
Increasing material cost *c* reduces the number of waves, while the attack rate generally increases with c. Panel a again depicts attack rate as a function of the material cost, and Panels b-e sample trajectories for different material cost values. At very low material costs, a small infection level is enough to trigger the behavioral change, and once the population adopts NPIs, relational costs (i.e. conformity) ensure it is maintained even with no infections (Panel b). Increased material cost (specifically, *c > θ*) means that after I drops to low levels conformity cannot maintain the NPI. Abandonment of the NPI sets the stage for repeated waves of infection and behavioral change. Initially the infection waves are small and numerous as the NPI adopted early in the wave due to relatively low cost (starting at three waves for these parameters; Panel c). As the material cost increases further, the NPI is both adopted later and abandoned sooner, which makes the waves bigger, and therefore fewer (Panels d and e). Eventually, if the cost is high enough, NPI is ever adopted, and the attack rate reaches its baseline at the far right side of panel a. Unless otherwise stated, the parameter values are taken from Table 1.

The constant *θ*, the weighting of relational utility or relational cost, can have varying effects on the attack rate. There are essentially three qualitative regimes from low to high *θ*: the existence of behavioural waves, locked in NPI usage once initiated, and no NPI usage. In the first regime, we observe a saw-tooth pattern in the attack rate for increasing relational cost. To understand this we considered a simplified scenario in which *ϵ* ≫ 1 and thus the population rapidly adopts or abandons NPI usage. In such a scenario, if the size of the susceptible population at the exit wave *S*_*x*_ is greater than 1*/R*_0_, then increasing *θ* increases the attack rate. On the other hand, if *S*_*x*_ *<* 1*/R*_0_ increasing *θ* can decrease the attack rate (though this is necessary but not sufficient condition; see Appendix A.4). In the second regime, the attack rate precipitously drops and there is a linear increase in the attack rate for further increasing *θ*. In this regime, only a single sustained behavioural response is induced, as can be observed in the time series of Figure 5e. The relational cost is sufficiently large that once NPI’s are used, relational utility is sufficient to promote its use even when infections subside. However, increasing *θ* after this regime has begun only delays the onset of NPI usage and therefore only increases the attack rate. In the third and final regime, *θ* is too high for the NPI to be adopted at all, and thus the attack rate is the same as for a non-behavioural model. Figure 5f depicts the best *θ* (i.e. the *θ* that minimizes the attack rate for a given *R*_0_). If *R*_0_ is too low (approximately *R*_0_ *<* 2), behavioural waves cannot be generated. Once they can be generated the best *θ* value increases as *R*_0_ increases, but saturates at *θ* ≈ 0.025. The minimum attack rate decreases even as *R*_0_ increases until *R*_0_ ≈ 4, after which further increases in *R*_0_ increase the minimum attack rate.

**Figure 5:**
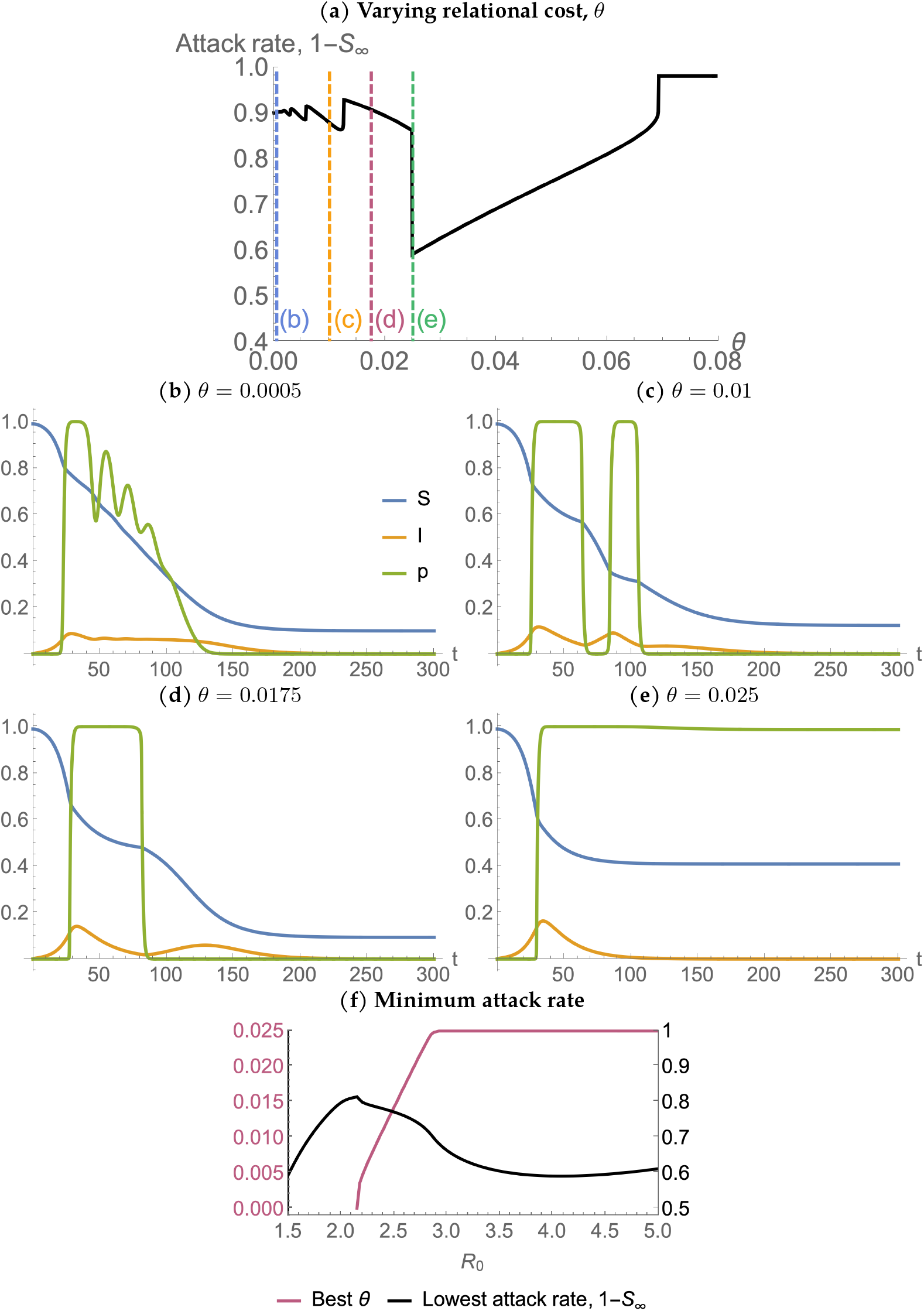
The effect of relational cost *θ* on the attack rate. Panel a shows that there are three regimes of *θ*’s impact on the attack rate: a reverse saw-tooth pattern, increasing attack rate, and a constant attack rate. As in the previous figures Panels b-e depict sample time trajectories for different regimes. For panel (f), we vary *β*_0_ so as to change *R*_0_. Unless otherwise stated, the parameter values are taken from Table 1.

## 4 Discussion

Physical and biological systems are frequently driven by human behaviour, which can be driven by what other people do (Martínez et al., 2021) as well as by group dynamics (Drury et al., 2021). Here, we have extended a canonical epidemiological model to incorporate human behaviour driven by risk of infection, personal NPI cost, efficacy of NPI’s, and social pressure of conformity. Our main finding is that the attack rate changes non-monotonically as a function of several different parameters.

First, we confirm a recent observation by Qiu et al. (2022) that attack rate displays a discontinuous non-monotonicity when increasing the transmission rate. At an initial threshold transmission rate *β*_0_, the attack rate suddenly drops due to the epidemic wave triggering behavioral change. After this initial threshold, the attack rate increases. In contrast to Qiu et al. (2022), however, we show that there can be additional thresholds where the number of epidemic and behavioral waves change, creating multiple distinct jumps, both up and down, in the attack rate as a function of transmission rate. This difference arises because we consider a slightly different model of behavioral change: in our model, individuals combine material and relational utilities and have a single threshold for adopting mask wearing, whereas Qiu et al. (2022) consider separate thresholds for fear of infection (corresponding to our material utility) and peer pressure (corresponding to our relational utility). In their model, once the entire population adopts the NPI behavior (their second tipping point), the peer pressure threshold ensures it’s permanently maintained even if infection levels go to zero, while in our model individuals will gradually return to the baseline behavior, provided the relational utility cost is less than the material utility cost. These results point to the additional complexity that can arise from the coupled epidemic and social dynamics.

Previous models have also considered individuals’ awareness of the epidemic Funk et al. (2009, 2010); Eksin et al. (2017, 2019); Weitz et al. (2020). The main difference between these models and ours is the inclusion of the relational utility and conformism in our behavioral spread model. These models therefore don’t display the stickiness of the NPI behavior we observe. Other differences also exist: Weitz et al. (2020), for example, consider the death rate as driving the awareness and the resulting reduction in transmission due to behaviour rather than the current number of infectious. This probably was a better description of the early stages of the COVID-19 epidemic in the US, when testing was scarce, and behavioral responses significantly lagged actual spread. They also incorporate “fatigue” where individuals get tired of the NPI, which can explain why mobility recovered in the US in late Spring 2020 despite high death rates. Weitz et al. (2020) also consider short and long term awareness of death rates; the latter can create some stickines in the NPI behavior. An interesting research question going forward is how longer-term changes in risk and relational preferences interact with each other.

One of our most surprising new results is that the efficacy of the NPI behavior also has a non-monotonic effect on the attach rate. Specifically, we show that the attack rate as a function of the efficacy of the NPI can have a saw-tooth pattern. Underlying this pattern is an interesting trade-off in increasing the NPI efficacy. The higher the NPI efficacy, the earlier the population reacts to the epidemic and the later it returns to normal, thereby increasing the duration of NPI adoption. This effect on its own reduces the size of the exit wave of the pandemic. However, NPI efficacy also decreases the transmission rate during the period it is being used thus leaving a larger pool of susceptibles and thereby a larger exit wave and overshoot. This suggests that intermediate level of NPI efficacy can be optimal. Alternatively, it suggests that policy interventions that encourage NPI adoption in the exit wave specifically to reduce overshoot can be effective in reducing final epidemic size.

We also observe interesting effects on epidemic dynamics and attack rates when varying the relational cost *θ* and material cost *c*. The former is due to the “stickiness” of the norm. Depending on the size of the exit wave and the basic reproductive number at that point, increasing the relational cost can be either beneficial or potentially harmful in impacting the attack rate. On one extreme, low relational costs lead to plateaus in infection rates and “flatten the curve” while having relatively small effect on final epidemic size. On the other hand, high enough relational costs can lock in the NPI once it is adopted (consider Japanese mask-wearing as an example of such locked in behaviour (Burgess and Horii, 2012)). However, the higher this cost, the more adoption is delayed, which increases the initial wave. At the extreme, NPIs might never get adopted due to conformism to the status quo. This suggests intermediate levels of relational costs might be optimal. For material costs, we find that, as might be expected, cheaper NPIs are better, as they get adopted faster and last longer. Increasing material costs introduces new waves and generally increased the attack rate. Overall, these results suggest that policies that reduce the material cost of NPIs as much as possible are beneficial, while for other parameters, the effects of increasing or decreasing them depends on the context, and there might be large jumps in the final epidemic size.

There are several factors that constitute both limitations of this study and opportunities for further research. We assumed individuals have homogeneous contact rates, and homogeneous costs. In reality, individuals belong to different communities with different costs and benefits, as well as different contact patterns. Such heterogeneity can, for example, be reflected in mask use (English and Li, 2021). In the appendix (Appendix B) we explore a model with two types of individuals with different material or relational costs. The results from this model are largely similar to our base model. On the other hand, heterogeneity between individuals can extend farther than simply the material costs, and might involve political views (Baxter-King et al., 2022), underlying differences in social norms, beliefs, or group identity (Wu and Huber, 2021). Smaldino and Jones (2021) explore a model with two groups of individuals which differ in both disease transmission and behavior adoption from within and outside their groups. They find that depending on the degree of homophily and outgroup aversion for adopting NPIs, one of the groups might experience a larger epidemic than otherwise, and each group might experience different epidemic waves. While Smaldino and Jones (2021) model behavioral change as a simple contagion process, these results likely will carry over to our norm-based behavioral model.

Another assumption we made is that individuals know the true level of NPI adoption of others. Though we explored the case with small positive or negative biases in these beliefs and found no large impact on the qualitative results (Appendix C), further exploration of the role of inaccurate beliefs in this model could be useful. In a previous compartmental model of norm adoption, it was found that such inaccurate beliefs about population behavior could substantially impact the emergence of a social norm (Morsky and Akçay, 2021). In the context of NPIs, biases in beliefs about population behavior can arise through visibility or salience bias (Han et al., 2019): for instance, people going out dining are more visible than people staying in, which can induce lower perception of NPI behaviors, and delay the onset of NPI adoption. However, some NPIs such as mask wearing will be less subject to such a bias. In addition to biases about population behavior, there might also be inaccurate beliefs about the material costs of infection and/or NPIs that can quicken or delay adoption and decay of NPIs.

Finally, we have not considered vaccination in this model. Fu et al. (2017) demonstrated that vaccination behavior is not random with respect to network degree and exposure to others who are vaccinated. This creates a second contagion process that can alter the dynamics of the epidemic, where the relative rates and network structure under which vaccination and the infection spread will determine the outcome. On some level, a transmission-reducing vaccine could be seen as analogous to an NPI but once adopted, its persistence does not depend on population level of vaccination. On the other hand, as the COVID-19 pandemic showed, vaccination can also be limited by supply factors and the willingness of individuals to receive it. Modeling these factors in conjunction with NPIs will add more realism to models of coupled socio-epidemiological dynamics, and likely add more wrinkles to the complex relationship between attack rate and the epidemiological parameters we present here.

## Data Availability

The code to run the numerical simulations and make figures is available at https://github.com/bmorsky/norms-and-pandemics.

## A Impact of varying parameters on attack rate for a single behavioural response

Consider the case where there is a single behavioural response and that it is immediate when initiated and that there is no exposed compartment (infected individuals are immediately infectious). The response occurs at threshold frequency of infections

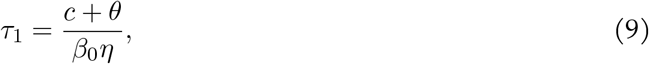

and stops at threshold frequency of infections

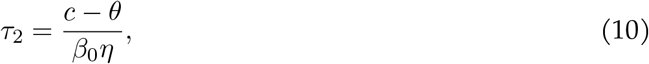

assuming *c > θ*. The final size calculation from time *t* = 0 to the point of the first threshold is

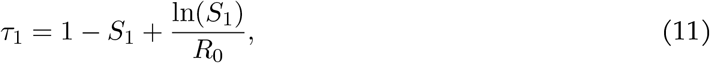

where *S*_1_ is the frequency of susceptibles at this threshold and *R*_0_ = *β*_0_*/γ*. We can do a similar calculation for the second threshold:

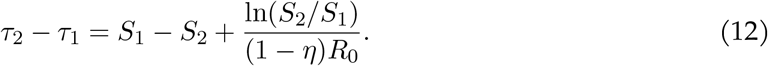

Finally, after this threshold (and assuming only one behavioural response) we have

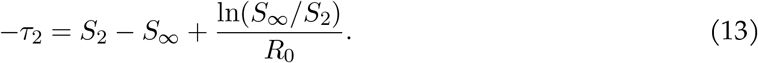

### A.1 Varying efficacy of the NPI *η*

Considering the first threshold, we have

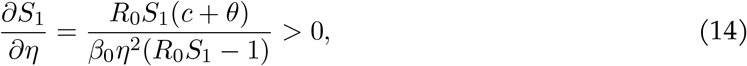

since *R*_0_*S*_1_ *>* 1. At the second threshold we have

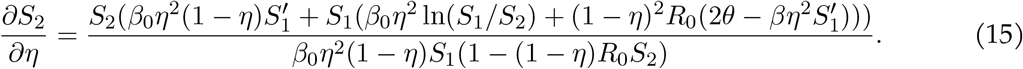

Finally, for 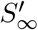 we have

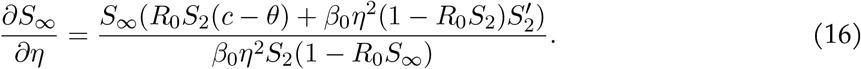

And, this is positive if *R*_0_*S*_2_ *<* 1, i.e. the epidemic continues to decline once the behaviour is stopped. *R*_0_*S*_2_ *>* 1 is a necessary but not sufficient condition for ∂*S*_∞_*/*∂*η <* 0: we must also have that 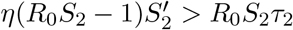.

### A.2 Varying the transmission rate *β*_0_

Considering the first threshold, we have

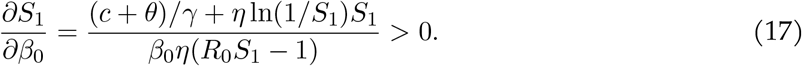

At the second threshold we have

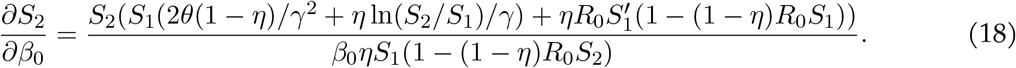

Finally, for ∂*S*_∞_*/*∂*β*_0_ we have

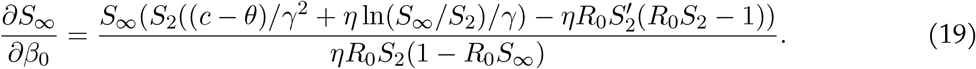

### A.3 Varying material cost *c*

Considering the first threshold, we have

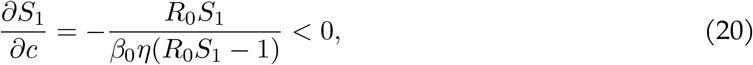

since if the threshold is going to be met, *R*_0_*S*_1_ *>* 1 (i.e. we can’t be on the other side of the wave of infections). Similarly, we know that (1 − *η*)*R*_0_*S*_2_ *<* 1. At the second threshold we have

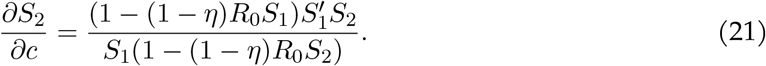

This is positive if (1 − *η*)*R*_0_*S*_1_ *>* 1 and negative if (1 − *η*)*R*_0_*S*_1_ *<* 1. Subbing these equations into ∂*S*_∞_*/*∂*c* gives

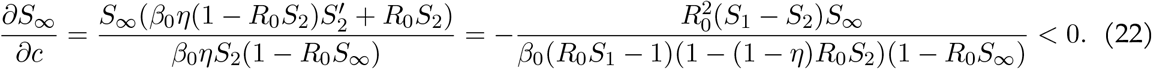

Thus, increasing *c* will only increase the attack rate 1 − *S*_∞_ assuming there is only one behavioural response wave.

### A.4 Varying the relational cost *θ*

Considering the first threshold, we have

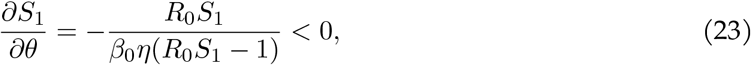

since *R*_0_*S*_1_ *>* 1. At the second threshold we have

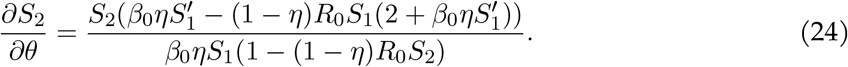

Finally, with respect to the final frequency of susceptibles we have

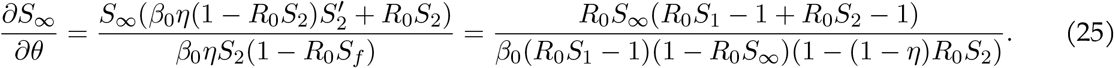

If *R*_0_*S*_2_ *>* 1, then ∂*S*_∞_*/*∂*θ >* 0. *R*_0_*S*_2_ *<* 1 is a necessary but not sufficient condition for ∂*S*_∞_*/*∂*θ <* 0.

## B Two type model

Another fundamental component of disease spread is heterogeneity within the population. It is well-established that heterogeneity in contact patterns (Bansal et al., 2007), age structure (Keeling, 1999), and individual susceptiblity (Colombo et al., 2020; Gomes et al., 2020) can dramatically affect infectious disease dynamics and the final outcome. Here, however, we incorporate behavioral heterogeneity in responding to a disease. In the case of COVID-19, for example, although elderly and immunocompromised individuals can be more susceptible to disease, they can also reduce their risk by taking appropriate precautions. Further, individual risk perception may not be accurate, and can be determined by one’s social environment. As such, the calculation of epidemiological quantities such as herd immunity levels and *R*_0_ are non-trivial, and can vary as the population is unevenly infected and changes its behaviour in response to the information and signalling they observe.

Since population heterogeneity can be an important factor in the spread of disease, we consider a variation of the above model where there are two types of individuals that are well-mixed with one another. Here the heterogeneity is in behaviour. They differ in the costs they experience when adopting the NPI. For example, one subset of the population may face higher material costs to social distancing than the other due to their type of work or lifestyle, which would make them relatively resistant to adopt the NPI. Sub-populations may differ in their sensitivity to social conformity to adopt the NPI, and hence *θ* could vary.

Consider two different sub-populations that are well mixed with one another, yet have have different costs. The equations for this model are:

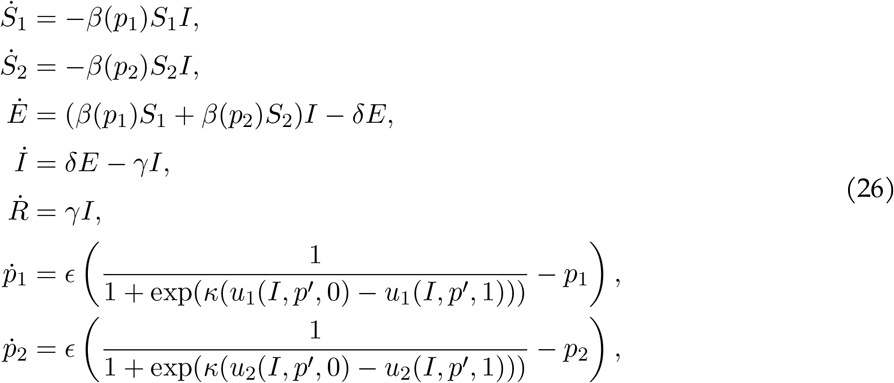

where *p*′ is the NPI usage individuals feel relational utility relative to. When they compare their behaviour to the total average behaviour, then 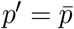, and when they compare it only to their type *p*′ = *p*_*j*_ for type *j*. The former we denote non-homophilous imitation and the second homophilous imitation. Utility for type *j* is thus:

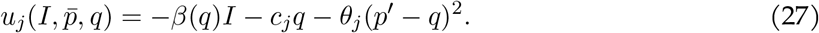

We consider either *c*_*j*_ ≠ *c*_*k*_ and *θ*_*j*_ = *θ*_*k*′_, or *c*_*j*_ = *c*_*k*_ and *θ*_*j*_ ≠ *θ*_*k*_.

Here we report the results for the two type population where material or relational costs differ. We consider the case where half of the population is of each type, and initially 0.005 of each type is exposed to the disease. Consider first the case where type 1 has a lower material cost to adopt the NPI than type 2, *c*_1_ = 0.02 *<* 0.04 = *c*_2_, and relational cost is the same *θ*_1_ = *θ*_2_ = 0.01. Figure 6 depicts this scenario when imitation is non-homophilous (panels a, b and c) and homophilous (panels d, e and f). The results are qualitatively similar to the single type model.

**Figure 6:**
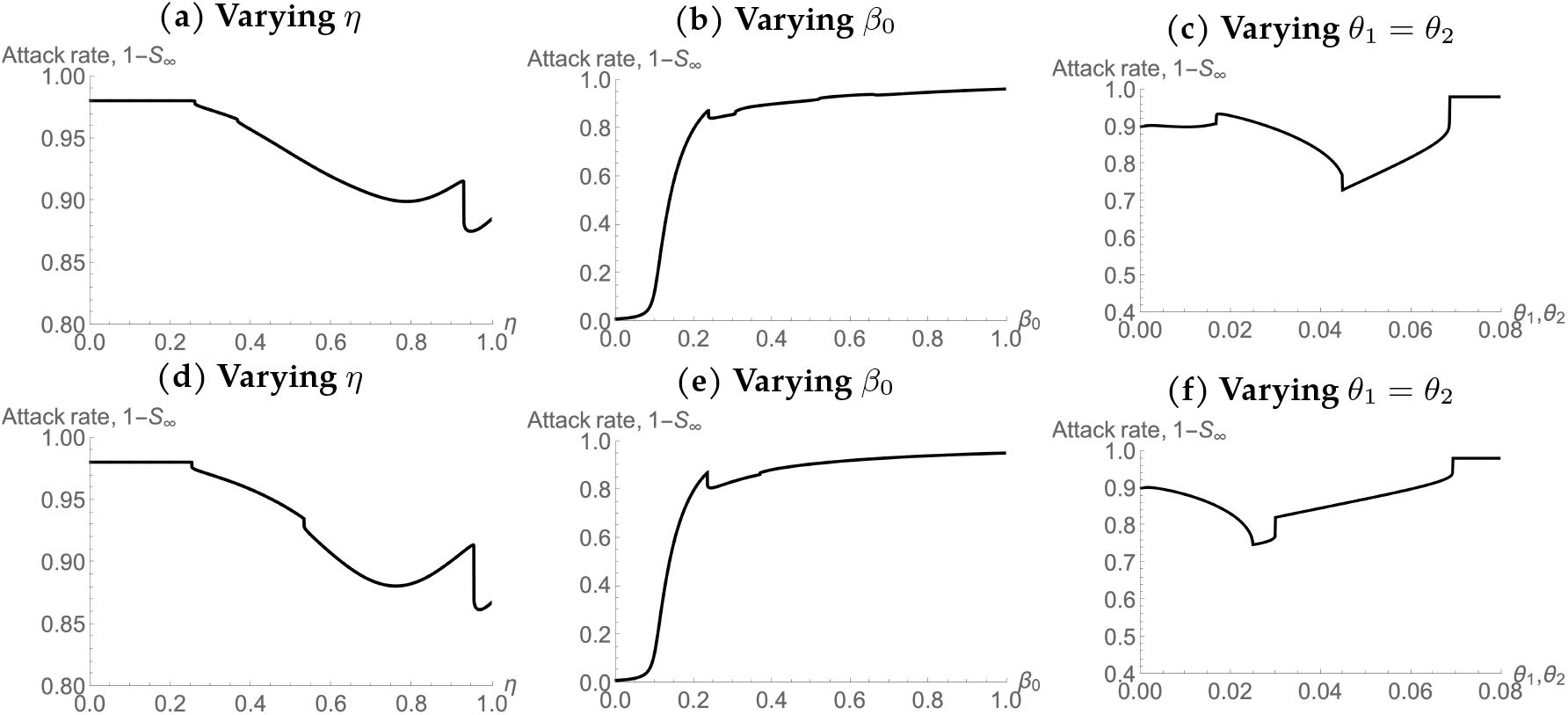
Attack rates as a function of different parameter values for the two-type scenario. Here *θ*_1_ = 0.02, *c*_2_ = 0.04, and *θ*_1_ = *θ*_2_. In the top row, imitation is non-homophilous, and in the bottom it is homophilous.

Now consider the case where type 1 has a lower relational cost to adopt the NPI than type 2: *θ*_1_ = 0.01 *<* 0.02 = *θ*_2_, and relational cost is the same *c*_1_ = *c*_2_ = 0.02. Figure 7 depicts this scenario when imitation is non-homophilous (panels a, b and c) and homophilous (panels d, e and f). The results are qualitatively similar to the single type model. The primary difference is that there are more jumps in the curves.

**Figure 7:**
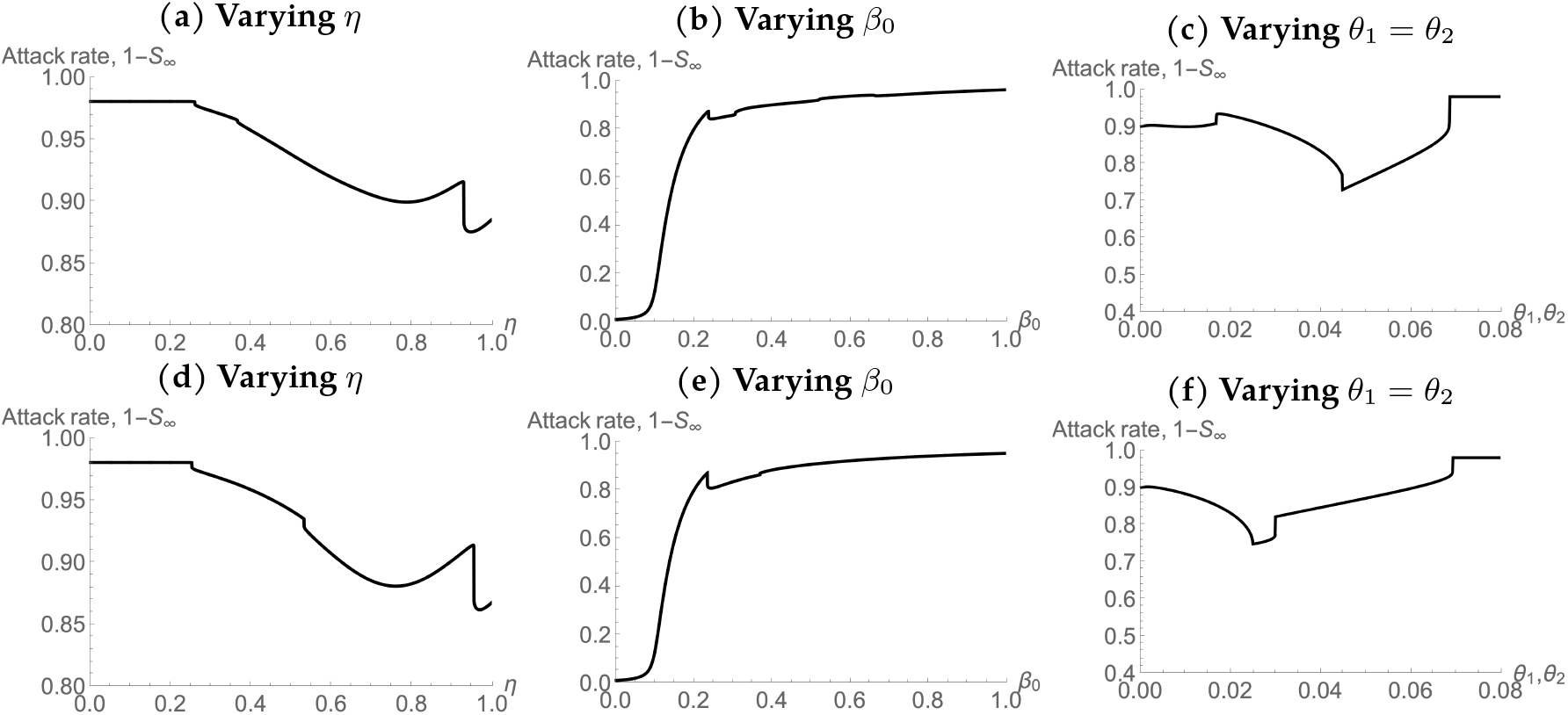
Attack rates as a function of different parameter values for the two-type scenario. Here *θ*_1_ = 0.02, and *θ*_2_ = 0.04. In the top row, imitation is non-homophilous, and in the bottom it is homophilous.

## C Biases in perceptions of norm compliance

Here we explore the case where the perception of NPI usage is positively or negatively biased. We do this by replacing *p* in the utility functions with 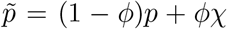, where *ϕ* ∈ [0, 1] is the degree of the bias and *χ* = 0 for negative bias and *χ* = 1 for positive bias. Figure 8 depicts the case for both types of biases with 20% bias (i.e. *ϕ* = 0.2). In general, the results have much of the similar results as in the non-biased case.

**Figure 8:**
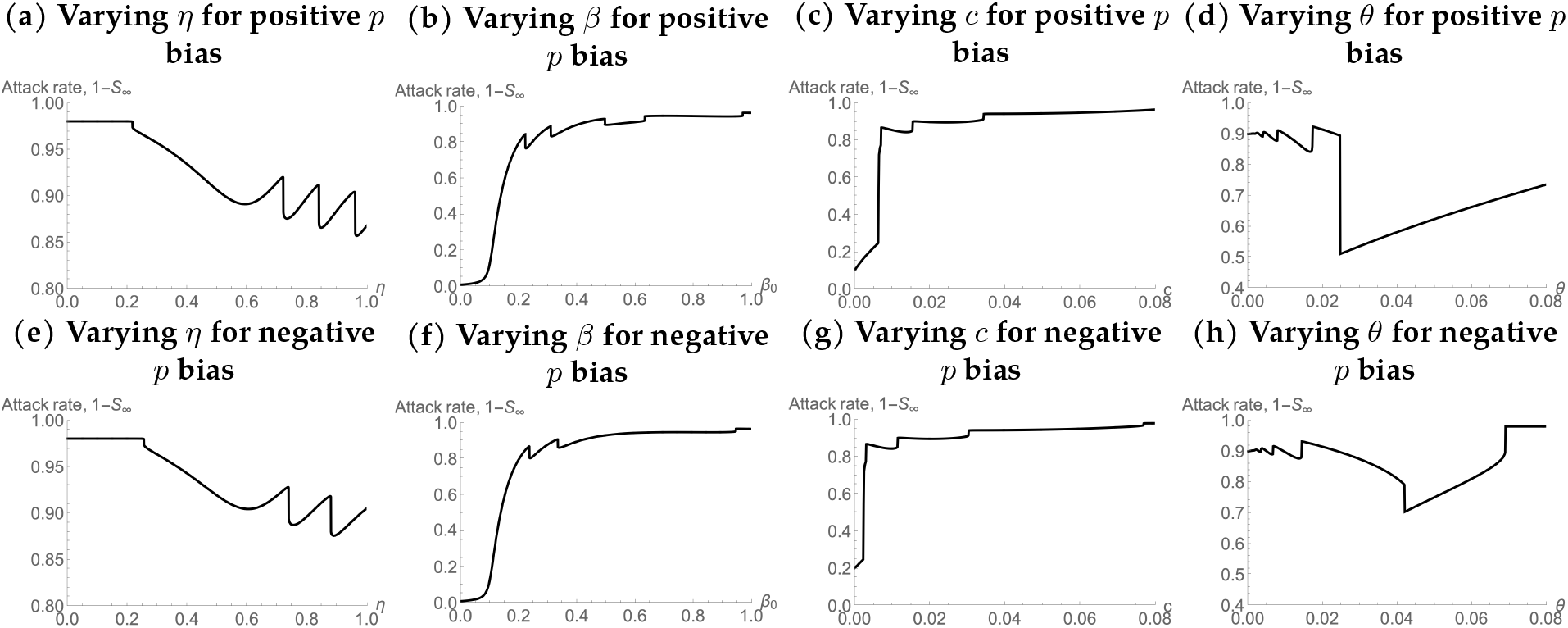
Attack rates for varying parameters with positive *p* bias (top row) and negative *p* bias (bottom row).

